# The effect of prehabilitation on health resource use and one-year survival: The Active Together Service

**DOI:** 10.1101/2025.09.01.25334844

**Authors:** Kerry Rosenthal, Nik Kudiersky, Gabriella Frith, Gail Phillips, Carol Keen, Daniel Howdon, Shaun Barratt, Diana M Greenfield, Gary Mills, Anna Myers, Liam Humphreys, Robert Copeland

**Author notes:** Corresponding author – Kerry Rosenthal, Advanced Wellbeing Research Centre, Sheffield Hallam University, Sheffield, S9 3TU, United Kingdom.

## Abstract

**Introduction:** Research suggests that multi-modal prehabilitation can improve quality of life and clinical outcomes. There is limited evidence, however, on the effect of prehabilitation on hospital resource use.

**Materials and Methods:** Patients receiving surgery for colorectal, lung or upper gastrointestinal cancer between January 2022 and March 2024, were referred to a multi-modal prehabilitation service (Active Together). Patients who declined to participate in Active Together and historical patient data (2017-2021) were used as comparator groups. Outcome measures were length of hospital stay, length of critical care stay, the total number of days spent in hospital as a readmission in the 90 days following surgery, and one-year survival rate.

**Results:** 305 patients completed prehabilitation, 96 patients declined to join the service, and 869 patients were included in the historical dataset. Active Together colorectal patients spent less time in critical care than historical colorectal patients (0.9 vs 1.2 days, p = 0.011). Whereas Active Together lung patients spent longer in critical care than historical lung patients (2.5 vs 1.7 days, p < 0.001). One-year survival rate was greater in Active Together patients compared to the declined group (95% vs 85%, p= 0.013), and not significantly different from the historical group (95% vs 92%, p= 0.14). The probability of the prehabilitation being cost saving was 58%, 60%, and 59% for colorectal, lung and upper gastrointestinal patients, respectively.

**Conclusion:** Prehabilitation has a limited effect on healthcare resource use and may improve one-year survival. Further research with large sample sizes is required to confirm small differences.

## 1. Introduction

Surgeries for colorectal, lung or upper gastrointestinal (GI) cancers are invasive, and it can take patients many months, if at all, to return to their pre-surgery functional ability [1]. There is also a high risk of surgery-related complications, which can lead to increased mortality and morbidity [2,3]. Prehabilitation aims to optimise patient resilience prior to surgery, improve postoperative outcomes, and facilitate recovery and increasingly involves a multi-modal intervention combining physical activity, dietetic and psychological support [4,5]. There is growing evidence that prehabilitation can improve clinical outcomes and quality of life [6,7]. Additionally, prehabilitation may improve patient eligibility for surgery [8-10], allowing patients to access treatments otherwise unavailable to them. Despite evidenced benefits, prehabilitation is not standard care for cancer patients across the UK.

The length of stay post-surgery is often used as an indication of the patient’s recovery and treatment costs. Reducing length of stay is part of the business case used to justify the investment in prehabilitation, however, it is a highly variable measure, as many factors can lead to a longer length of stay, such as comorbidities, clinical judgement, and day of admission [11]. A previous meta-analysis for exercise interventions prior to surgery for lung cancer patients significantly reduced length of stay [12]. Similarly, a meta-analysis found a reduction in length of stay for abdominal cancer patients who received prehabilitation compared to controls [13]. The individual studies did not find significant differences [14-16], indicating the need for large sample sizes to identify changes in this measure.

A recent study on prehabilitation prior to colorectal surgery showed an average reduction in hospital stay (0.91 days) and a cost saving of €140 per patient [17]. Two systematic reviews of prehabilitation cost analyses show that generally, prehabilitation for surgery tends to be cost-effective; however, they caution for the risk of publication bias [18,19].

Previous evidence suggests that prehabilitation improves disease-free survival in colorectal cancer patients [20,21]. However, the evidence in this area for other cancers remains limited [12,22]. A three-year rehabilitation programme for patients with colorectal cancer resulted in significantly improved 8-year survival rates (90.3% survival compared to 83.2%), although limited differences were seen after one-year.

Active Together is a prehabilitation and rehabilitation service in Sheffield, UK that provides multimodal support for patients undergoing treatment for lung, colorectal or upper gastrointestinal cancer [23,24]. The service is integrated into the NHS pathway, and all eligible patients are referred to the service. This paper considers the impact Active Together on healthcare resource use and one-year survival in a real-world clinical setting.

## 2. Methods

### 2.1 Ethics

The Health Research Authority reviewed the evaluation protocol [24] and concluded that formal ethical approval was not required because there was no change in patient care. The service evaluation was approved by the Clinical Effectiveness Unit at Sheffield Teaching Hospitals NHS Foundation Trust (Ref 11115—May 19, 2022). Evaluation data was pseudonymised prior to analysis to ensure patient confidentiality. Patients attending Active Together provided informed consent to share hospital data for the evaluation. Patients that did not attend Active Together were included if they had not opted out using the NHS National Data Opt-Out Service.

### 2.2 Prehabilitation

Patients receiving curative treatment for colorectal, lung or upper gastrointestinal cancer were referred to Active Together at the point of diagnosis. Active Together provided personalised physical activity, dietetic and psychological support, depending on patients’ level of need, which was determined by a range of factors including existing comorbidities and baseline level of fitness. Patients with a higher level of need received specialist support such as 1-to-1 sessions with a physiotherapist, dietician or clinical psychologist, and participated in group exercise classes, whereas low-need patients received home exercise programmes and online resources. All Active Together patients were offered support throughout their cancer treatment and up to six months of rehabilitation (three months restorative and three months supportive) [24]. Detailed descriptions of the needs assessment and support offered are available elsewhere [23]. The length of prehabilitation varied depending on the treatment wait times. Between January 2022 and March 2024, 425 patients received surgery for colorectal, lung or upper gastrointestinal cancer after attending prehabilitation at Active Together. Patients who only received other forms of treatment (e.g. radical radiotherapy) but not surgery were not included in this analysis. 104 patients were excluded from further analysis, as they had not received at least 2 weeks of prehabilitation.

### 2.3 Comparator Groups

As Active Together is an NHS clinical service and not a research trial, a concurrent control group was unavailable. Patients who were referred to Active Together but declined to participate in the service (N = 96) were therefore used as a comparator group. An historical dataset comprising of 3427 surgeries performed between January 2017 and December 2021 was obtained to use as an additional comparator group. The use of historical data as a comparator group is an established approach used in single-arm research trials [25].

### 2.4 Clinical outcomes

The length of stay in hospital and length of stay in critical care after surgery were obtained from hospital records. Additionally, the total number of days spent in hospital as a readmission in the 90 days following surgery was included as a secondary outcome. Date of death was used to calculate one-year survival rate.

### 2.5 Outliers

The mean and standard deviation of the historical data were used to exclude length of stay outliers more than 3.29 standard deviations above the mean, as done previously [26]. This equated to a length of stay over 44 days. Five Active Together outliers and 47 Historical outliers were removed. There were no length of stay outliers in the Declined group.

### 2.6 Matched controls

To minimise bias related to variation in surgical complexity, Active Together patients were randomly matched to up to three historical controls [27], by procedure type and whether their tumour was malignant (N = 282) or benign (N = 23). Data was not available to match by disease progression or cancer stage; however, all patients were undergoing treatment with curative intent. A list of included surgical procedures is available in Appendix 1. A total of 11 Active Together patients with no appropriate match were removed from further analysis. The final sample sizes for each group are shown in Table 1.

**Table 1.** Sample sizes and characteristics for each tumour and prehabilitation group.

| Tumour group | Prehabilitation group | N | Mean Age (SD) | Male sex (%) |
| --- | --- | --- | --- | --- |
| Colorectal | Active Together | 162 | 68.6 (10.0) | 53.7 |
|  | Declined | 31 | 68.0 (10.5) | 51.6 |
|  | Historical | 463 | 66.8 (11.1) | 55.9 |
| Lung | Active Together | 81 | 67.8 (9.0) | 46.9 |
|  | Declined | 51 | 70.6 (8.2)* | 47.1 |
|  | Historical | 243 | 70.1 (9.0)* | 51.9 |
| Upper GI | Active Together | 62 | 65.1 (9.6) | 69.4 |
|  | Declined | 14 | 68.6 (7.9) | 78.6 |
|  | Historical | 163 | 65.2 (10.0) | 78.5 |
| Total | Active Together | 305 | 67.7 (9.7) | 55.1 |
|  | Declined | 96 | 69.5 (8.9) | 53.1 |
|  | Historical | 869 | 67.4 (10.5) | 59.0 |

\* Significantly different ( $P < 0.05$ ) from Active Together group.

### 2.7 Cost estimation

The estimated cost of the Active Together service was £712.86 per referred patient. This included support provided before and after surgery. This was calculated from the annual cost of running the service (including salary, premises, equipment, training, travel and miscellaneous costs) divided by the number of referrals received per year.

The hospital stay cost for surgical cancer treatment for each patient was calculated using the methods described by Arabadzhyan and colleagues [28]. HRG4+ National Costs Grouper was used to assign HRG codes to each hospital spell and associated costs drawn from the 2022-23 National Cost Collection data series.

### 2.8 Data analysis

T-test and Fishers’ exact tests were used to compare the age and sex, respectively, between the Active Together group to the Declined and Historical groups. A Spearman’s rank correlation was performed to test whether it was appropriate to include age as a covariate for length of stay analysis; however, there was no correlation between age and length of stay (rho = 0.003, p = 0.927).

The Mann-Whitney U test with a Benjamini-Hochberg correction for the two comparisons (to Declined and Historical groups) was used to compare healthcare resource use. P < 0.05 was considered statistically significant, and a Cohen’s d effect size over 0.2 was considered meaningful [29]. Both means and medians are presented. The data is not normally distributed; however, means are more clinically meaningful in this instance as they will reflect the total per-patient cost incurred by the hospital (including outliers).

A cost analysis was performed to determine whether Active Together was associated with increased overall treatment costs. Data was only available to compare to the Declined group. The analysis included the cost of the Active Together programme as well as the costs incurred during the initial hospital stay, including surgical procedures. Independent t-tests were used to compare the treatment costs between Active Together and Declined group for each tumour group. A probability of cost-saving is provided in line with conventional health economic methods [30], using 10,000 bootstrap samples. Where results show positive improvements in health outcomes, this can be considered to be a lower limit on the probability of cost-effectiveness (i.e., a conservative estimate).

One-year survival post-surgery was analysed using a Cox proportional hazards regression model. P < 0.05 was considered statistically significant.

## 3. Results and Discussion

### 3.1 Characteristics

The group characteristics were not significantly different between prehabilitation groups, apart from in the lung cancer patients, where Active Together patients were significantly younger than the declined or historical groups (Table 1).

### 3.2 Prehabilitation length

The average prehabilitation length (from first appointment with Active Together until their first treatment) and length of support before surgery is shown in Table 2.

**Table 2.** Mean (SD) Active Together prehabilitation length for each tumour group.

| Tumour Group | Mean prehabilitation length in days (SD) | Mean days from first appointment to surgery (SD) |
| --- | --- | --- |
| Colorectal | 47.7 (34.7) | 56.7 (52.7) |
| Lung | 26.3 (14.2) | 41.4 (30.8) |
| Upper GI | 33.8 (14.4) | 102.7 (43.0) |
| Total | 36.8 (22.8) | 62.2 (50.6) |

### 3.3 Reasons for declining

The reasons for declining Active Together given by the declined group are shown in Figure 1. The most common reasons for declining were ‘Not interested’ (N = 31) and ‘Self-managing’ (N = 28). These responses may reflect that patients do not believe they require structured support, or could indicate perceived barriers to participation, such as time constraints or a belief that existing self-care strategies are sufficient. Only two patients gave the reason ‘too unwell’, who were both lung cancer patients. Despite being too unwell to join the service, they did not have particularly long length of stays (9 and 3 days), and both survived over one year after their surgery.

**Figure 1.**
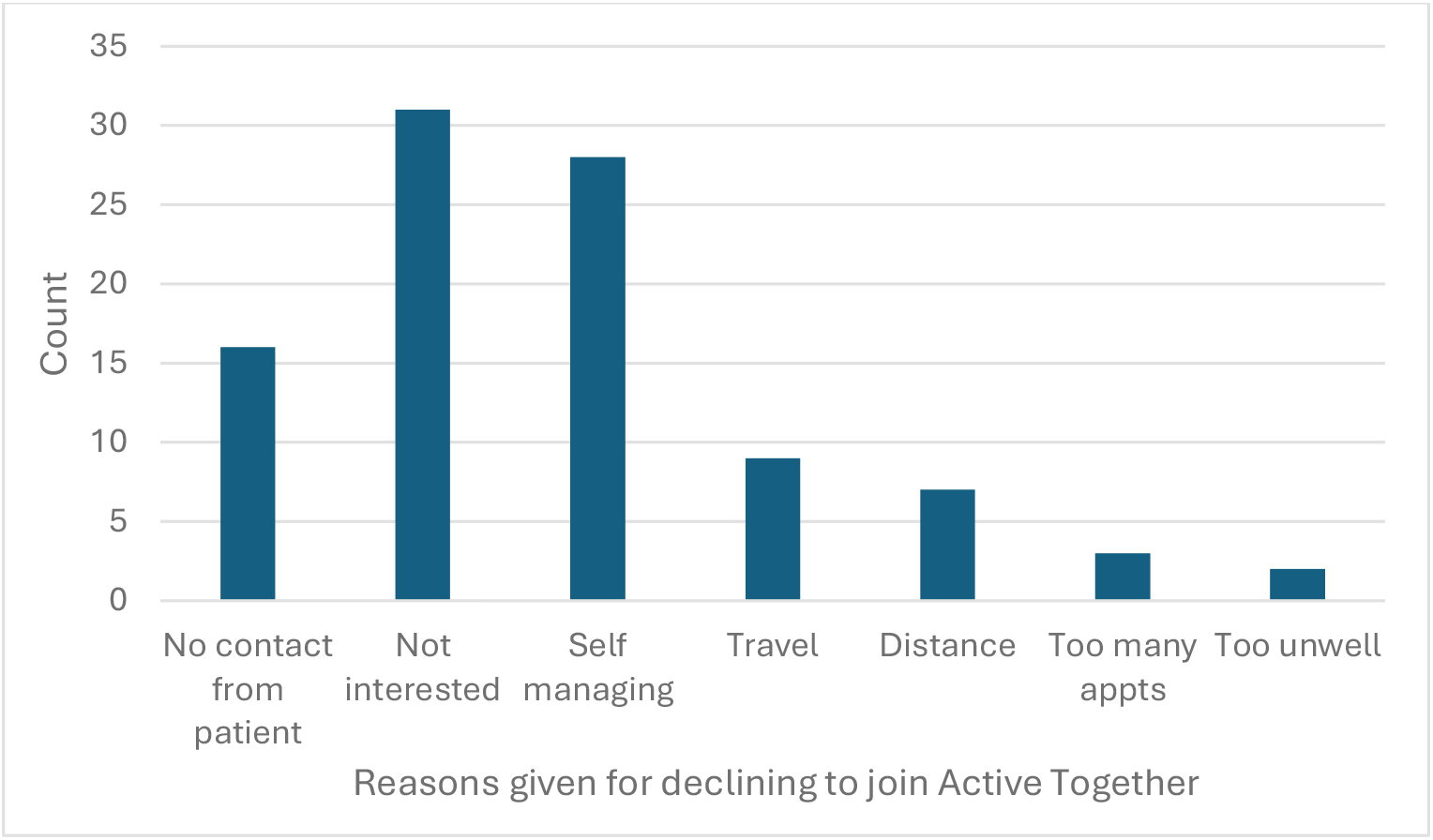
Reasons given by the ‘Declined’ group for not participating in the Active Together service.

### 3.4 Length of Stay

There were no significant differences in length of stay between the prehabilitation groups, overall or for the tumour sub-groups (Table 3). There was a meaningful improvement (as defined by an effect size > 0.2) in length of stay for the upper GI Active Together patients, with declined and historical patients staying 0.23 and 1.61 days longer, respectively. This represents a meaningful reduction in NHS resources; however, a larger dataset is required to confirm this trend. These results are similar to previous studies, which found nonsignificant reductions in length of stay after prehabilitation [14-16]. A similar service based in Greater Manchester, Prehab4Cancer, found significant reductions in length of stay after prehabilitation, for colorectal, lung, and oesophageal cancer patients [31]. Their non-prehabilitation groups had a longer mean length of stay compared to the Sheffield historical patient cohort, which could indicate a difference in regional hospital practice. Therefore, there may have been greater opportunity for improvement. In the Active Together analysis, all upper GI cancer groups had a shorter mean length of stay than the prehabilitation group in the Prehab4Cancer report. This demonstrates that the potential reductions in length of stay through prehabilitation may vary across hospitals, depending on current practice.

**Table 3.**
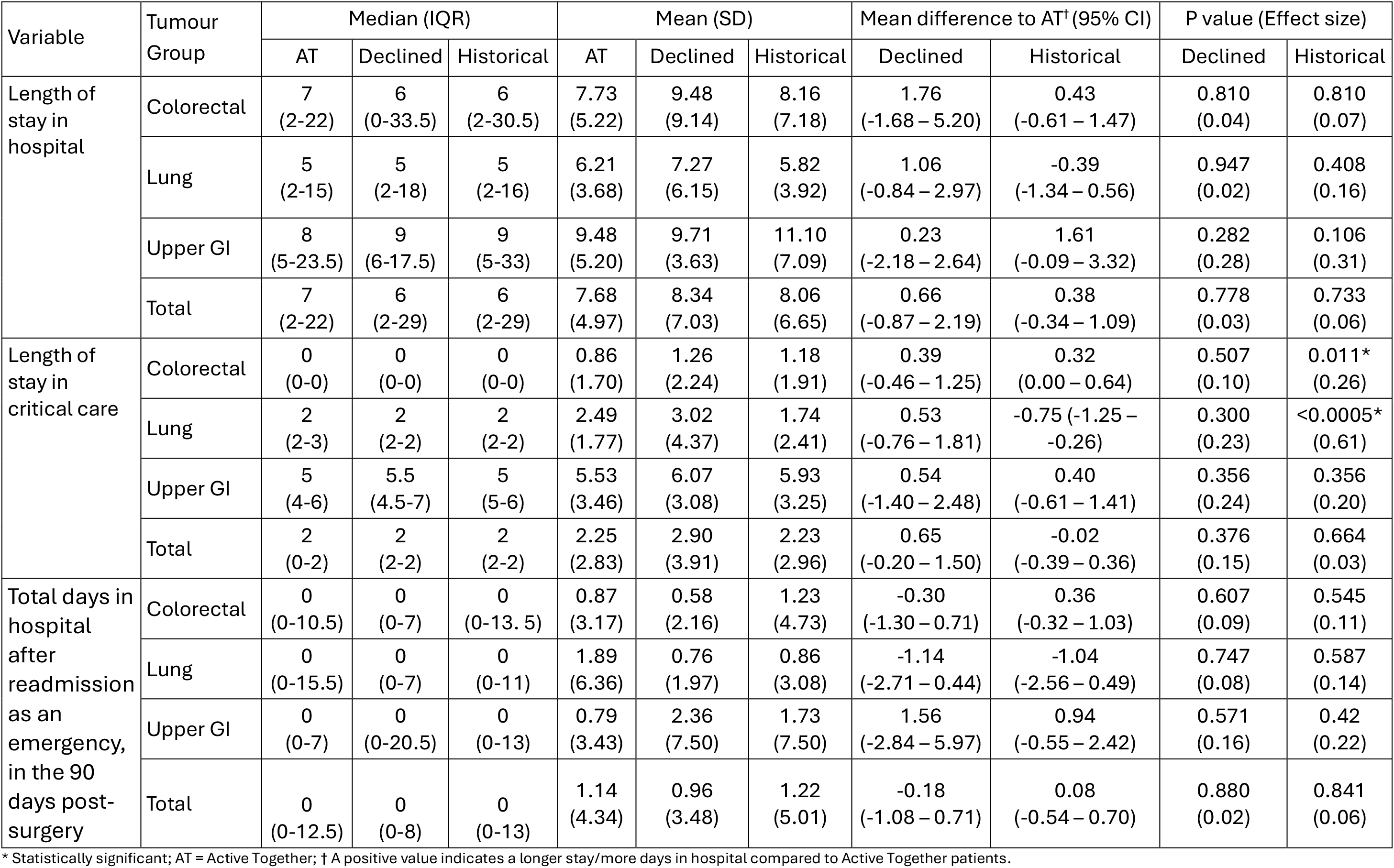
Hospital resource use after surgery for Active Together (AT), declined and historical patients.

Historical colorectal patients had a significantly longer (+0.32 days) stay in critical care than Active Together patients. Similarly, the Declined colorectal group had a longer stay in critical care than Active Together colorectal patients, although this was not statistically significant (Table 3). This is similar to the result Prehab4Cancer found for their colorectal cancer patients [31]. In this evaluation, there was a similar non-significant trend for upper GI patients.

Historical lung cancer patients spent significantly less time (−0.75 days) in critical care than Active Together patients. This difference may be partially explained by the inclusion of higher-risk patients in the Active Together cohort - individuals who may not have previously been offered surgery due to poor baseline fitness. In some cases, patients were referred to Active Together with the goal of becoming fit enough for surgery through prehabilitation. As a result, the Active Together group may have included patients at greater risk of postoperative complications, potentially requiring longer hospital stays. Prior to the inception of Active Together, these patients would not have received surgery and, therefore, would not be included in the Historical dataset. The number of patients that were not eligible for surgery at the start of prehabilitation was not systematically recorded; we are only aware of this anecdotally. Therefore, the effect on the final sample is uncertain. A previous study on prehabilitation for lung cancer patients, however, found that only 41% patients were fit for surgery before prehabilitation, whereas after prehabilitation, 76% were fit for surgery [10]. This suggests that almost 60% of patients who were ineligible for surgery, became eligible after prehabilitation.

### 3.5 Hospital Readmissions

There were no significant differences in the total number of days spent in hospital after an emergency readmission in the 90 days post-surgery (Table 3). This finding aligns with a previous meta-analysis which also found no significant difference in 30-day hospital readmissions between prehabilitation and control groups in patients undergoing abdominal cancer surgery [13]. As there are many possible reasons for emergency readmission, this measure may not be specific enough to capture any differences due to improved preoperative fitness.

### 3.6 Surgery Stay Cost

There were no significant differences between the total cost of hospital stay and prehabilitation for Active Together patients compared to the declined group (Table 4). For all three tumour groups, the mean cost was higher for the declined group than the Active Together group, with average cost savings of £260, £494, and £477 for colorectal, lung and upper GI cancer patients, respectively. This is similar to a previous study that showed a statistically non-significant cost-saving for colorectal cancer patients [17]. While not statistically significant, if these cost-savings are extrapolated to a national level, it could save the NHS £19 million per year, based on the number of lung and gastrointestinal cancer cases [32] and the percentage that receive surgery [33].

**Table 4.**
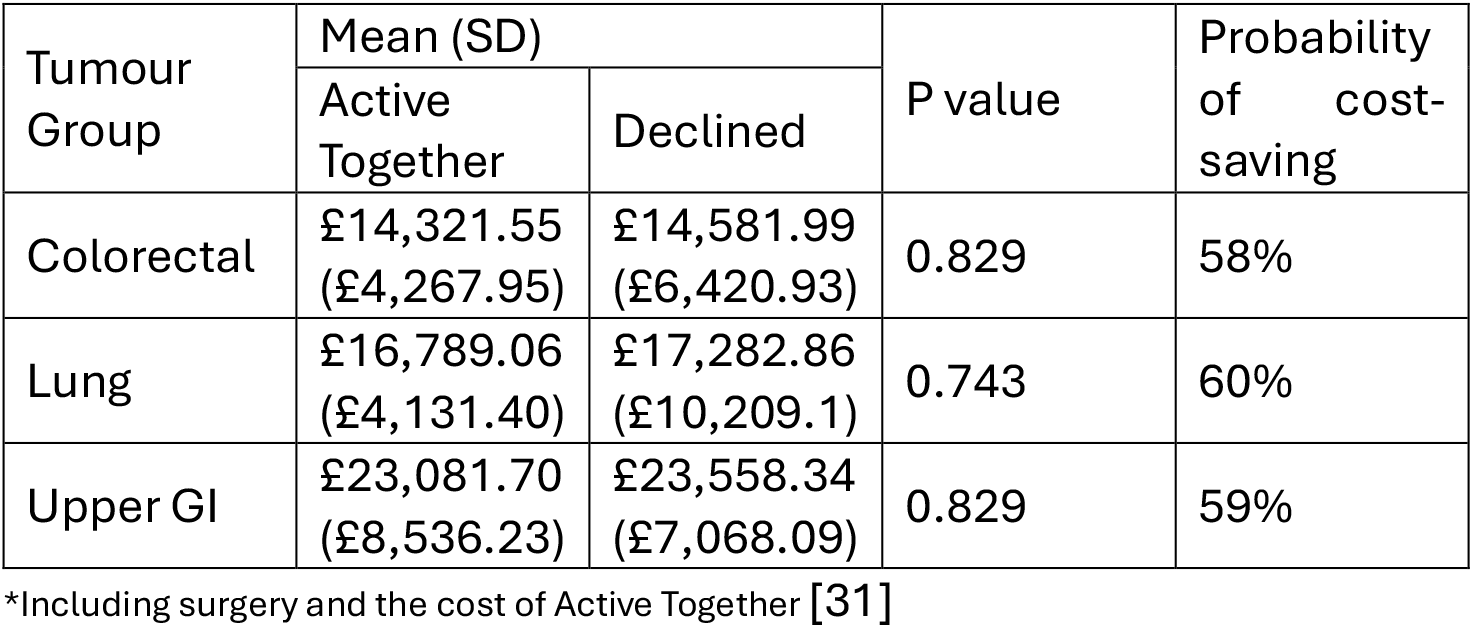
Total cost of treatment* for Active Together and Declined patients.

The probability of cost-saving was in all cases greater than 50%. Given the improvement in fitness and wellbeing outcomes exhibited across patients in the Active Together group [34], this can be interpreted as a lower limit (i.e., conservative estimate) on the probability of cost-effectiveness and give confidence that the intervention is more likely than not to be cost-effective, assuming the declined group is a representative sample.

The Active Together model stratifies patients based on their level of need. The actual cost of delivering support therefore varies between individuals. For the purpose of this evaluation, the same cost of Active Together (£712.86) has been allocated to each patient, as estimating the true cost would be very complex. This cost includes the whole Active Together pathway (prehabilitation, maintenance and rehabilitation), not just prehabilitation costs. The short-term cost savings would be greater, if only the cost of prehabilitation was included. There may be other long-term cost savings that are not captured by this analysis, such as reduced reliance on primary care services.

Provision based on needs may be the most cost-effective method, as those with higher initial needs will likely benefit the most, as suggested previously [18]. However, delivering support to these patients typically incurs higher costs, due to the requirement for closer supervision and more intensive, in-person support. As only Active Together patients underwent a needs assessment, it was not possible to compare the effectiveness of the intervention on sub-groups of high or low needs patients in this evaluation.

Active Together is a clinician-led service, which employs physiotherapists to provide physical activity support to patients with complex needs, alongside specialist fitness instructors for lower needs patients. In contrast, the leisure-sector led model, delivered without clinical supervision is more likely to exclude patients with more complex health conditions. This high-risk cancer population potentially have the most to gain from prehabilitation, as they face greater risks of treatment-related complications and functional decline. Additionally, a prehabilitation service for ovarian cancer patients based in Wales found that engagement improved when they moved to a physiotherapy-based service, rather than an exercise-referral service, suggesting that patients placed higher importance in a clinician-led service [35]. The creation of a service that is delivered predominantly by allied health professionals allows for the inclusion of the most complex and high-risk patients, and may improve overall engagement, but requires appropriate resource with cost implications. The optimal prehabilitation model is yet to be determined.

The type of surgery may also influence whether the intervention is cost-effective. It has been suggested that prehabilitation may be more cost-saving when the surgery type is particularly aggressive [36]. Larger sample sizes would allow for differences between surgery types and complexity to be examined.

### 3.7 One-Year Survival

For all tumour groups, the percentage of patients surviving one-year was highest in the Active Together group, compared to the Declined or Historical groups. This difference was significant for the upper GI group and, overall, compared to the declined group (Table 5). This provides promising evidence of the positive effect of prehabilitation and rehabilitation on overall survival; however, the historical comparisons may be confounded by improvements to cancer treatments. Similarly, declined groups may have been more unwell or from disadvantaged backgrounds, and may receive different treatment from healthcare professionals who were aware of whether patients are participating in the service. Longer term examinations of overall and disease-free survival are necessary to build on this area, across cancer types.

**Table 5.** Survival rates and hazard ratios from Cox regression model.

| Tumour Group | Statistic | Active Together | Declined | Historical |
| --- | --- | --- | --- | --- |
| Colorectal | 1-Year Survival<br>(95% CI) | 97%<br>(94% – 100%) | 86%<br>(72% – 100%) | 95%<br>(93% – 97%) |
|  | Hazard Ratio<br>(95% CI)* | 1 | 3.49<br>(0.78 – 15.6) | 2.15<br>(0.76 – 6.07) |
|  | P value* |  | 0.100 | 0.150 |
| Lung | 1-Year Survival<br>(95% CI) | 93%<br>(87% – 99%) | 89%<br>(80% – 99%) | 90%<br>(86% – 94%) |
|  | Hazard Ratio<br>(95% CI)* | 1 | 1.32<br>(0.48 – 3.63) | 1.13<br>(0.53 – 2.42) |
|  | P value* |  | 0.600 | 0.750 |
| Upper GI | 1-Year Survival<br>(95% CI) | 91%<br>(83% – 100%) | 68%<br>(45% – 100%) | 85%<br>(80% – 91%) |
|  | Hazard Ratio<br>(95% CI)* | 1 | 3.88<br>(1.39 – 10.8) | 1.31<br>(0.61 – 2.83) |
|  | P value* |  | 0.010 | 0.490 |
| Total | 1-Year Survival<br>(95% CI) | 95%<br>(92% – 98%) | 85%<br>(77% – 93%) | 92%<br>(90% – 94%) |
|  | Hazard Ratio<br>(95% CI)* | 1 | 2.29<br>(1.19 – 4.40) | 1.43<br>(0.89 – 2.30) |
|  | P value* |  | 0.013 | 0.140 |

\* Compared to Active Together patients

## Conclusions

Patients that received prehabilitation in a real-world setting did not have higher overall healthcare costs compared to those that declined the service and had a higher one-year survival rate. Combined with the positive impact on patient outcomes, these results build upon previous research demonstrating the positive impact of integrating prehabilitation services like Active Together into standard pre-surgical care pathways for cancer patients, particularly for colorectal and upper GI cancers where small but meaningful improvements were observed. Larger-scale evaluations and clinical comparator studies are needed to confirm the positive trends on length of stay and one-year survival and understand the impact on access to surgery.

## Data Availability

Data are not publicly available.

## Credit author statement

KR: methodology, formal analysis, data curation, writing (original draft). NK: methodology, writing (review and editing). GF: methodology, writing (review and editing). GP: project administration. CK: methodology, investigation, resources. DH: methodology, formal analysis, writing (review and editing). SB: software, data curation. DG: conceptualization, writing (review and editing). GM: conceptualization, writing (review and editing). AM: methodology. LH: conceptualization, methodology, writing (review and editing), funding acquisition. RC: conceptualization; methodology, supervision, funding acquisition, resources.

